# Anterior nasal versus nasal mid-turbinate sampling for a SARS-CoV-2 antigen-detecting rapid test: does localisation or professional collection matter?

**DOI:** 10.1101/2021.02.09.21251274

**Authors:** Olga Nikolai, Chiara Rohardt, Frank Tobian, Andrea Junge, Victor M. Corman, Terry C. Jones, Mary Gaeddert, Federica Lainati, Jilian A. Sacks, Joachim Seybold, Frank P. Mockenhaupt, Claudia M. Denkinger, Andreas K. Lindner

## Abstract

**Objectives:** The aim of this diagnostic accuracy study was direct comparison of two different nasal sampling methods for an antigen-based rapid diagnostic test (Ag-RDT) that detects severe acute respiratory syndrome coronavirus 2 (SARS-CoV-2). Furthermore, the accuracy and feasibility of self-sampling was evaluated.

**Methods:** This manufacturer-independent, prospective diagnostic accuracy study, compared professional anterior nasal (AN) and nasal mid-turbinate (NMT) sampling for a WHO-listed SARS-CoV-2 Ag-RDT. A second group of participants collected a NMT sample themselves and underwent a professional nasopharyngeal swab for comparison. The reference standard was real-time polymerase chain reaction (RT-PCR) using combined oro-/nasopharyngeal sampling. Individuals with high suspicion of SARS-CoV-2 infection were tested. Sensitivity, specificity, and percent agreement were calculated. Self-sampling was observed without intervention. Feasibility was evaluated by observer and participant questionnaires.

**Results:** Among 132 symptomatic adults, both professional AN- and NMT-sampling yielded a sensitivity of 86.1% (31/36 RT-PCR positives detected; 95%CI: 71.3-93.9) and a specificity of 100.0% (95%CI: 95.7-100). The positive percent agreement (PPA) was 100% (95%CI: 89.0-100). Among 96 additional adults, self NMT- and professional NP-sampling yielded an identical sensitivity of 91.2% (31/34; 95%CI 77.0-97.0). Specificity was 98.4% (95%CI: 91.4-99.9) with NMT- and 100.0% (95%CI: 94.2-100) with NP-sampling. The PPA was 96.8% (95%CI: 83.8-99.8). Most participants (85.3%) considered self-sampling as easy to perform.

**Conclusion:** Professional AN- and NMT-sampling are of equivalent accuracy for an Ag-RDT in ambulatory symptomatic adults. Participants were able to reliably perform the NMT-sampling themselves, following written and illustrated instructions. Nasal self-sampling will likely facilitate scaling of SARS-CoV-2 antigen testing.

## Introduction

Due to their short turn-around time and ease-of-use, antigen-detecting rapid diagnostic tests (Ag-RDTs) enable new testing strategies for severe acute respiratory syndrome coronavirus 2 (SARS-CoV-2) [1, 2]. Currently, most SARS-CoV-2 Ag-RDTs require nasopharyngeal (NP) sampling, which is frequently perceived as uncomfortable and requires healthcare professionals, thus limiting scale-up. Nasal sampling could enable self-sampling and increase acceptability.

The term nasal sampling is often not used uniformly. The US Centers for Disease Control and Prevention (CDC) differentiates anterior nasal (AN) and nasal mid-turbinate (NMT) sampling [3]. Recent studies have demonstrated the equivalence of NMT-compared to NP-sampling for a WHO-listed SARS-CoV-2 Ag-RDT and the feasibility of self-sampling [4-6]. Recently, the CDC has added home/self NMT-sampling as an acceptable alternative to professional NP-sampling in their guidance for SARS-CoV-2 testing [3]. AN-sampling is easier and more convenient than NMT-sampling, but Ag-RDT performance with AN-sampling has not been evaluated.

The objective of this prospective diagnostic accuracy study was a head-to-head comparison of professional AN- and NMT-sampling for a WHO-listed SARS-CoV-2 Ag-RDT. Furthermore, the accuracy and feasibility of self NMT-sampling was evaluated.

## Methods

### Study design and participants

This was a manufacturer-independent, prospective diagnostic accuracy study comparing two different nasal sampling methods for an Ag-RDT. From a first group of participants, professionally-collected AN and NMT samples were taken. In a second group, each participant self-collected a NMT sample and underwent a professional NP swab. All samples were tested by Ag-RDT performed by a professional (Figure 1). The reference standard was real-time polymerase chain reaction (RT-PCR) using a combined oro-/nasopharyngeal (OP/NP) sample as described previously [6].

**Figure 1.**
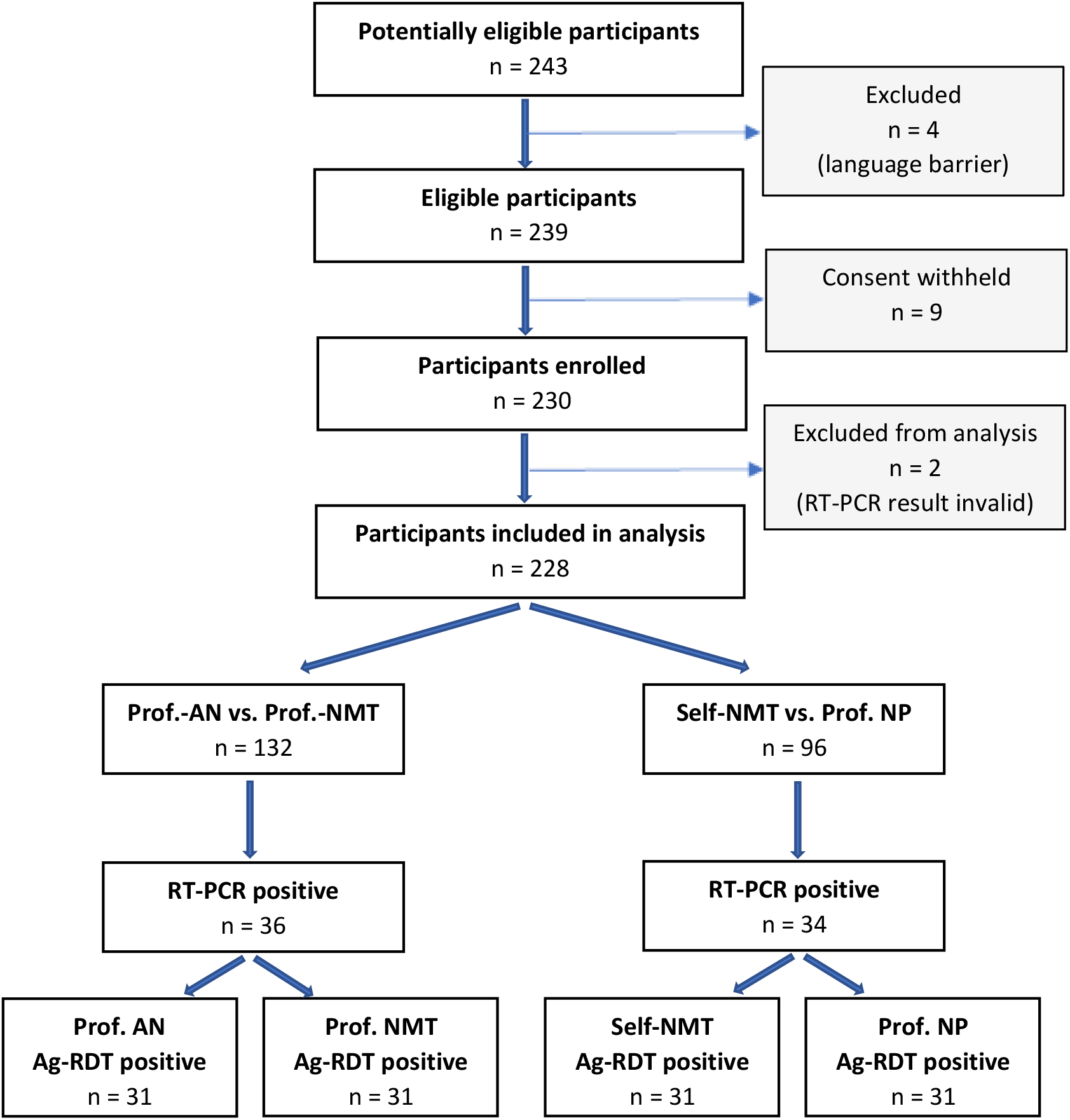
Study flow diagram Prof.-AN,professional anterior nasal sampling; prof.-NMT, professional nasal mid-turbinate sampling; self-NMT, self NMT-sampling; prof-NP, professional nasopharyngeal sampling; Ag-RDT, antigen-based rapid diagnostic test; RT-PCR, real-time polymerase chain reaction

The study took place at the ambulatory SARS-CoV-2 testing facility of Charité University Hospital between 30 November 2020 and 18 January 2021. Participants eligible for inclusion were adults with high clinical suspicion of SARS-CoV-2 infection. For self-sampling, a minimum CEFR (Common European Framework of Reference) language level of B2 (upper intermediate) in German or English was required. Participants were consecutively enrolled, according to laboratory capacity.

### Index test Ag-RDT

The Ag-RDT evaluated was the STANDARD Q COVID-19 Ag Test (SD Biosensor, Inc. Gyeonggi-do, Korea), which is also distributed by Roche in Europe [7]. While the test is commercially available as a NP-sampling kit, the nasal-sampling kit (used for NMT and AN) is currently for research use only. Differences between the swabs and the procedures of the two test kits have previously been described [4].

### Sampling methods

Participants were asked to blow their nose once before sampling. Professional AN- and NMT-sampling followed the CDC guidance for SARS-CoV-2 testing [3]. For AN-sampling, the tip of a swab was inserted into the nose vertically 1 to 1.5 cm and rotated against the nasal walls for 15 seconds in both nostrils. For NMT-sampling, while tilting the head back (70°) the swab was inserted horizontally (parallel to the palate) into both nostrils for about 2 cm until resistance occurred, and then rotated 4 times against the nasal walls. Among consecutive participants, the sequence of AN- and NMT-sampling was alternated, followed by OP/NP-sampling for RT-PCR.

Participants who underwent NMT self-sampling received written and illustrated instructions in German or English. For NMT self-collection, a timing of 15 seconds was specified in addition to the minimum of 4 rotations. Procedures were observed without answering questions or providing corrections. NMT self-sampling (both nostrils) was followed by professional NP-sampling (through one nostril) for Ag-RDTs and combined OP/NP-sampling (other nostril) for RT-PCR.

The Ag-RDTs were performed directly after sampling at point-of-care by study physicians with a semi-quantitative visual read-out of the test band as described in a prior study [6]. User acceptability and feasibility of self-sampling were assessed by observer and patient questionnaires [6].

## Results

### Participants

After the exclusion of 2 participants with invalid RT-PCR result, 132 participants with professional AN-versus NMT-sampling, and 96 who underwent self NMT-sampling versus professional NP-sampling were included (Figure 1). Average age was 34.6 years (Standard Deviation [SD] 11.7) with 46.7% females and 20.3% having comorbidities. On the day of testing, 97.4% of participants had one or more symptoms consistent with SARS-CoV-2 infection. Average duration of symptoms at the time of presentation was 3.4 days (SD 3.0). Among participants performing self-sampling, 48 (50.5%) had a prior swab for SARS-CoV-2 been collected, and 50 (52.6%) had a higher education degree (Supplementary Table 1).

### Professional AN-versus NMT-sampling

Among 132 participants, 36 (27.3%) were RT-PCR-positive for SARS-CoV-2. Professional AN- and NMT-sampling both yielded a sensitivity of 86.1% (31/36 RT-PCR positives detected; 95%CI: 71.3-93.9) and a specificity of 100.0% (95%CI: 95.7-100) compared to RT-PCR (Table 1, Supplementary Table 2). The positive percent agreement was 100% (95%CI: 89.0-100). There was perfect (100%) inter-reader agreement on results.

**Table 1:**
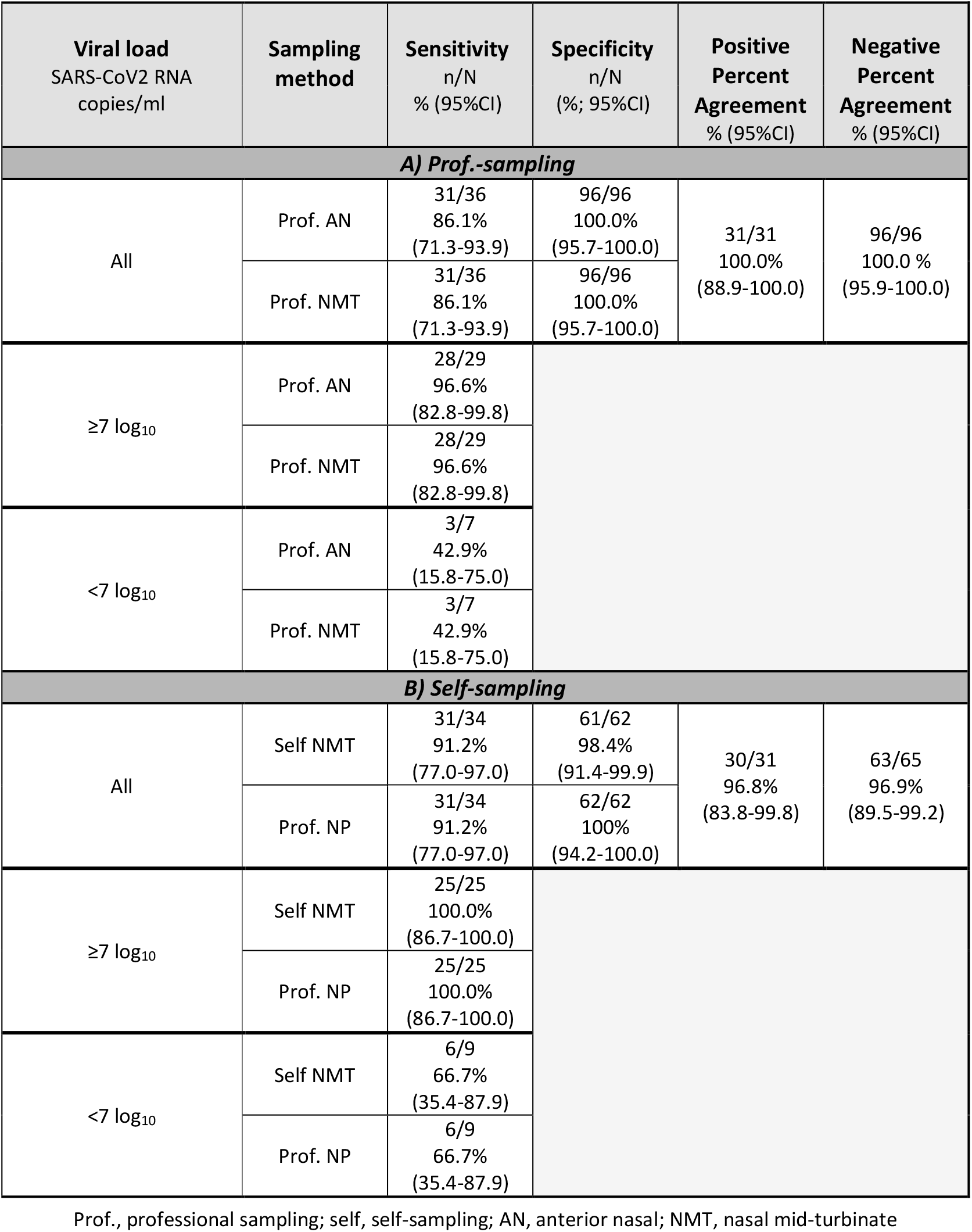
Sensitivity, specificity, and percent agreements of A) professional AN- versus professional NMT-sampling, and B) self NMT- versus professional NP-sampling. The results are also differentiated by high and low viral load (≥ /< 7 log_10_ SARS-CoV2 RNA copies/ml).

### Self NMT-sampling versus professional NP-sampling

Among 96 participants, 34 (35.4%) were RT-PCR-positive. Self NMT- and professional NP-sampling yielded an identical sensitivity of 91.2% (31/34; 95%CI: 77.0-97.0). Specificity was 98.4% (95%CI: 91.4-99.9) with self NMT-sampling and 100.0% (95%CI: 94.2-100) with NP-sampling (Table 1, Supplementary Table 3). The positive percent agreement was 96.8% (95%CI: 83.8-99.8). A third reader was necessary to agree on the interpretation of one NMT-result, which was ultimately considered negative, but turned out to be false negative based on a positive RT-PCR being result.

### Feasibility of self NMT-sampling

Deviations of self NMT-sampling included a more vertically-directed angle for sampling (n=13), incorrect depth (n=4 too superficial, n=10 too deep), and reduced swabbing intensity (regarding duration n=28, rotations n=12, and rubbing n=36). Three participants performed only unilateral NMT-sampling (Supplementary Table 3 and 4). On a scale from 1 (easy) to 5 (difficult), 81 (85.3%) participants stated that self NMT-sampling was easy to perform (scale 1 or 2); 13 (13.7%) found it medium easy/difficult (scale 3), and 1 (1.1%) rather difficult (scale 4). Twelve participants suggested that a mark on the swab to guide insertion depth would facilitate self-sampling.

## Discussion

Among symptomatic outpatients, the sensitivities in detecting SARS-CoV-2 with an Ag-RDT were identical with professional AN- and NMT-sampling (86.1% overall; and 96.6% at high viral load). Furthermore, self NMT-sampling yielded the same sensitivity as professional NP-sampling (91.2% overall; and 100% at high viral load). Thus, our data suggests that AN-sampling is a suitable alternative to NMT- or NP-sampling.

AN- and NMT-sampling protocols may overlap in practice and deviate in details [8]. Participants in this study blew their nose once, on the theoretical assumption that this may increase the virus concentration in the sampling region. Also a timing of 15 seconds was specified for self NMT-sampling in contrast to other protocols [3]. The sensitivities obtained in the present study are slightly higher compared to recent studies for the same Ag-RDT at this testing facility, most likely due to more patients presenting by chance with high viral load [4-6].

The strengths of the study are the rigorous standardized sampling methods, two independent blinded readers, and an additional semi-quantitative assessment of the Ag-RDT results. A limitation of the study is that it was performed in a single centre. Patients who performed self-sampling were rather young and educated, half of whom already had experienced professional sample collection for SARS-CoV-2.

The clinical usefulness of nasal swabs has been demonstrated and acknowledged for SARS-CoV-2 RT-NPCR, including self-sampling [8-10], and evidence for Ag-RDTs is growing [4-6, 11, 12]. With written and illustrated instructions, patients were able to easily perform NMT-sampling. Procedural sampling-deviations might be reduced by video instructions. Nasal self-sampling will allow scaling of antigen testing. Considering the diagnostic equivalence, the more convenient self AN-sampling should allow an even broader use.

## Data Availability

All raw data and analysis code are available upon request to the corresponding author.

## Author contributions

AKL, ON, and CMD designed the study and developed standard operating procedures. ON and CR implemented the study design and performed the laboratory work. AJ enrolled participants and supported the laboratory work. ON, CR, and AKL led the writing of the manuscript. FPM and JS coordinated and supervised the study site. FT and MG led the data analysis. FL provided technical advice. VMC and TCJ were responsible for PCR testing and contributed to the interpretation of the data. JAS supported the study design setup. All authors have reviewed the manuscript.

## Acknowledgements

Heike Rössig, Maximilian Gertler, Susen Burock, Franka Kausch, Mia Wintel, Julian Bernhard, Niklas Krug, Elisabeth Linzbach, Melanie Bothmann, Zümrüt Tuncer, Stefanie Lunow, Beate Zimmer, Astrid Barrera Pesek, Sabrina Pein, Verena Haack, Oliver Deckwart, Birgit Zittlau.

## Data availability

All raw data and analysis code are available upon request to the corresponding author.

