## Supplementary Table 1 for "Anterior nasal versus nasal mid-turbinate sampling for a SARS-CoV-2 antigen-detecting rapid test: does localisation or professional collection matter?"

**Supplementary Table 1.** Characteristics of participants undergoing self-sampling

| | | |
|---|---|---|
| **Swab for COVID-19 taken in the past** N=95 | n (%) | 48 (50.5%) |
| **General school qualification** N=90<br>- lower secondary school<br>- intermediate secondary school<br>- upper secondary school | n (%) | <br>2 (2.2%)<br>17 (18.9%)<br>71 (78.9%) |
| **Highest further education** N=95<br>- none<br>- vocational degree<br>- higher education degree | n (%) | <br>16 (16.8%)<br>35 (36.8%)<br>50 (52.6%) |
| **Profession** N=95<br>- health professionals<br>- teaching professionals<br>- technicians and associate professionals<br>- other professionals<br>- clerical support, service, sales workers<br>- craft and related trades workers<br>- trainees, students<br>- others | n (%) | <br>5 (5.3%)<br>18 (18.9%)<br>7 (7.4%)<br>19 (20.0%)<br>15 (15.8%)<br>3 (3.2%)<br>20 (21.1%)<br>8 (8.4%) |
| **Instruction in non-native language** N=96 | n (%) | 16 (16.7%) |
| **CEFR language level (English/German)** N=16<br>- B2 (upper intermediate)<br>- C1 (advanced)<br>- C2 (mastery) | n (%) | <br>5 (31.3 %)<br>3 (18.8%)<br>8 (50%) |

COVID-19, Coronavirus disease 2019; CEFR language level, Common European Framework of Reference for Languages
