## Supplementary Table 2 for "Anterior nasal versus nasal mid-turbinate sampling for a SARS-CoV-2 antigen-detecting rapid test: does localisation or professional collection matter?"

**Supplementary Table 2.** Ag-RDT results with a professional-collected NMT swab and AN swab in RT-PCR positive patients, together with RT-PCR cycle threshold (Ct) values, viral load (in descending order), and duration of symptoms. The positive percent agreement between NMT and AN samples on Ag-RDT, and the respective sensitivities compared to RT-PCR are shown.

| No. | Ag-RDT | | RT-PCR (OP/NP) | | Days of Symptoms |
|---|---|---|---|---|---|
| | Prof. NMT | Prof. AN | Ct value | Viral load[1] | |
| 1 | pos. (+++) | pos. (+++) | 12.65 | 10.22[2] | 1 |
| 2 | pos. (+++) | pos. (+++) | 14.63 | 9.63[2] | 2 |
| 3 | pos. (+++) | pos. (+++) | 15.19 | 9.46[2] | 1 |
| 4 | pos. (+++) | pos. (+++) | 15.25 | 9.44[2] | 2 |
| 5 | pos. (+++) | pos. (+++) | 15.42 | 9.39[2] | 3 |
| 6 | pos. (+++) | pos. (+++) | 15.66 | 9.32[2] | 4 |
| 7 | pos. (+++) | pos. (+++) | 15.71 | 9.31[2] | 1 |
| 8 | pos. (+++) | pos. (+++) | 15.74 | 9.30[2] | 3 |
| 9 | pos. (+++) | pos. (+++) | 15.89 | 9.25[2] | 1 |
| 10 | pos. (+++) | pos. (+++) | 16.58 | 9.05[2] | 2 |
| 11 | pos. (+++) | pos. (+++) | 16.73 | 9.00[2] | 2 |
| 12 | pos. (+++) | pos. (+++) | 16.80 | 8.98[2] | 2 |
| 13 | pos. (+++) | pos. (+++) | 17.08 | 8.90[2] | 4 |
| 14 | pos. (+++) | pos. (+++) | 17.43 | 8.80[2] | 2 |
| 15 | pos. (+++) | pos. (+++) | 17.56 | 8.76[2] | 4 |
| 16 | pos. (+++) | pos. (+++) | 17.63 | 8.74[2] | 2 |
| 17 | pos. (+++) | pos. (+++) | 20.73 | 8.58[3] | 2 |
| 18 | pos. (+++) | pos. (++) | 18.46 | 8.49[2] | 3 |
| 19 | pos. (+++) | pos. (+++) | 18.68 | 8.42[2] | 2 |
| 20 | pos. (+++) | pos. (+++) | 21.30 | 8.41[3] | 1 |
| 21 | pos. (+++) | pos. (+++) | 21.33 | 8.40[3] | 2 |
| 22 | pos. (+++) | pos. (+++) | 18.81 | 8.39[2] | 7 |
| 23 | pos. (+++) | pos. (+++) | 21.75 | 8.28[3] | 4 |
| 24 | pos. (+++) | pos. (+++) | 19.47 | 8.19[2] | 2 |
| 25 | pos. (+++) | pos. (+++) | 22.27 | 8.12[3] | 7 |
| 26 | pos. (+++) | pos. (+++) | 20.33 | 7.93[2] | 4 |
| 27 | neg. | neg. | 20.59 | 7.86[2] | 1 |
| 28 | pos. (+++) | pos. (+++) | 20.88 | 7.77[2] | 1 |
| 29 | pos. (++) | pos. (+) | 20.95 | 7.75[2] | 6 |
| 30 | pos. (+++) | pos. (+++) | 26.81 | 6.78[3] | 1 |
| 31 | neg. | neg. | 25.15 | 6.50[2] | 3 |
| 32 | neg. | neg. | 29.28 | 6.05[3] | 1 |
| 33 | pos. (++) | pos. (++) | 27.18 | 5.90[2] | 5 |
| 34 | neg. | neg. | 27.28 | 5.87[2] | 1 |
| 35 | pos. (++) | pos. (++) | 29.74 | 5.14[2] | 5 |
| 36 | neg. | neg. | 31.38 | 4.65[2] | 5 |
| | **Sensitivity** 31/36 (86.1%) | **Sensitivity** 31/36 (86.1%) | | | |
| | **Positive percent agreement** 100.0% | | | | |

[1] $log_{10}$ SARS-CoV2 RNA copies/ml
[2] TibMolbiol assay, E-gene target
[3] Roche Cobas SARS-CoV-2 assay (E-gene, T2 target)

Prof., professional sampling; self, self-sampling; AN, anterior nasal; NMT, nasal mid-turbinate; Ag-RDT, Antigen-based rapid diagnostic tool; RT-PCR, Real-time polymerase chain reaction; OP, oropharyngeal; NP, nasopharyngeal; Ct, cycle threshold; No, Number; pos. (+), weak positive; pos. (++), positive; pos. (+++), strong positive; neg., negative
