## Supplementary Table 3 for "Anterior nasal versus nasal mid-turbinate sampling for a SARS-CoV-2 antigen-detecting rapid test: does localisation or professional collection matter?"

**Supplementary Table 3** Antigen-detecting RDT results with a self-collected NMT swab and a professional-collected NP swab in RT-PCR positive patients, as well as Ct-values, viral load (in descending order), and duration of symptoms per patient. The positive percent agreement between NMT and NP samples on Ag-RDT, and the respective sensitivities compared to RT-PCR are shown.

| No. | Ag-RDT | | RT-PCR (OP/NP) | | Days of Symptoms | Part of self-sampling with relevant deviation |
|---|---|---|---|---|---|---|
| | Self NMT | Prof. NP | Ct value | Viral load[1] | | |
| 1 | pos. (+++) | pos. (+++) | 13.48 | 9.97[2] | 5 | rotation |
| 2 | pos. (+++) | pos. (+++) | 14.74 | 9.60[2] | 1 | |
| 3 | pos. (+++) | pos. (+++) | 14.97 | 9.53[2] | 3 | |
| 4 | pos. (+++) | pos. (+++) | 18.88 | 9.13[3] | 2 | |
| 5 | pos. (+++) | pos. (+++) | 16.41 | 9.10[2] | 2 | rubbing |
| 6 | pos. (+++) | pos. (+++) | 19.67 | 8.89[3] | 2 | rotation, rubbing |
| 7 | pos. (+++) | pos. (+++) | 17.13 | 8.89[2] | 2 | rotation, rubbing |
| 8 | pos. (+++) | pos. (+++) | 17.89 | 8.66[2] | 4 | |
| 9 | pos. (+++) | pos. (+++) | 20.58 | 8.62[3] | 2 | rubbing |
| 10 | pos. (+++) | pos. (+++) | 18.05 | 8.61[2] | 8 | |
| 11 | pos. (+++) | pos. (+++) | 18.08 | 8.60[2] | 2 | |
| 12 | pos. (+++) | pos. (+++) | 18.08 | 8.60[2] | 2 | rubbing |
| 13 | pos. (+++) | pos. (+++) | 20.71 | 8.59[3] | 1 | rotation, rubbing |
| 14 | pos. (+++) | pos. (+++) | 18.80 | 8.39[2] | 4 | |
| 15 | pos. (+++) | pos. (+++) | 21.64 | 8.31[3] | 4 | |
| 16 | pos. (+++) | pos. (+++) | 19.13 | 8.29[2] | 3 | rotation, rubbing |
| 17 | pos. (+) | pos. (++) | 22.74 | 7.98[3] | 4 | anterior head tilt |
| 18 | pos. (+++) | pos. (+++) | 22.85 | 7.95[3] | 1 | AN localisation |
| 19 | pos. (+++) | pos. (+++) | 20.28 | 7.95[2] | 4 | rotation, rubbing |
| 20 | pos. (+++) | pos. (+++) | 22.98 | 7.91[3] | 2 | rubbing |
| 21 | pos. (++) | pos. (+++) | 22.99 | 7.91[3] | 7 | |
| 22 | pos. (+++) | pos. (+++) | 21.83 | 7.49[2] | 2 | rotation |
| 23 | pos. (++) | pos. (+++) | 24.49 | 7.47[3] | 7 | unilateral |
| 24 | pos. (+) | pos. (++) | 21.92 | 7.46[2] | 1 | rotation, vertical angle |
| 25 | pos. (+++) | pos. (+++) | 22.76 | 7.21[2] | 5 | rotation |
| 26 | pos. (++) | pos. (+++) | 26.40 | 6.90[3] | 5 | rubbing |
| 27 | pos. (+++) | pos. (+++) | 26.59 | 6.85[3] | 6 | vertical angle |
| 28 | pos. (+++) | pos. (+++) | 27.13 | 6.69[3] | - | AN localisation, rubbing |
| 29 | neg. | neg. | 27.66 | 6.53[3] | 3 | rotation |
| 30 | pos. (+++) | pos. (+++) | 27.82 | 6.48[3] | 1 | vertical angle, rotation, rubbing |
| 31 | pos. (++) | pos. (+) | 25.46 | 6.41[2] | 9 | rotation |
| 32 | neg. | pos. (++) | 25.85 | 6.29[2] | 3 | rotation, rubbing |
| 33 | pos. (++) | neg. | 30.31 | 5.75[3] | 6 | |
| 34 | neg. | neg. | 30.42 | 4.93[2] | 20 | rubbing |
| | **Sensitivity** 31/34 (91.2%) | **Sensitivity** 31/34 (91.2%) | | | | |
| | **Positive percent agreement** 96.8% | | | | | |

[1] $log_{10}$ SARS-CoV2 RNA copies/ml
[2] TibMolbiol assay, E-gene target
[3] Roche Cobas SARS-CoV-2 assay (E-gene, T2 target)

Prof., professional sampling; self, self-sampling; AN, anterior nasal; NMT, nasal mid-turbinate; Ag-RDT, Antigen-based rapid diagnostic tool; RT-PCR, Real-time polymerase chain reaction; OP, oropharyngeal; NP, nasopharyngeal; Ct, cycle threshold; No, Number; pos. (+), weak positive; pos. (++), positive; pos. (+++), strong positive; neg., negative
