## Supplementary Table 4 for "Anterior nasal versus nasal mid-turbinate sampling for a SARS-CoV-2 antigen-detecting rapid test: does localisation or professional collection matter?"

**Supplementary Table 4.** The relevant steps of self NMT-sampling with numbers of participants who precisely followed the instructions, according to observations of study physicians.

| **Individual steps of self NMT-sampling**<br>N=96 | | **Participants following precisely instructions** |
|---|---|---|
| - removal of the swab from packing | n (%) | 80 (83.3%) |
| - swab only touched at handle | | 94 (97.9%) |
| - reclination of head | | 83 (86.5%) |
| - insertion about 2 cm | | 92 (95.8%) |
| - angle of the sampling direction | | 83 (86.5%) |
| - rotation for minimum 15 seconds | | 68 (70.8%) |
| - rotation minimum 4 times | | 84 (87.5%) |
| - rubbing against the nasal walls N=95 | | 59 (62.1%) |
| - bilateral sampling | | 93 (96.9%) |
